# The future of computational pathology: expectations regarding the anticipated role of artificial intelligence in pathology by 2030

**DOI:** 10.1101/2022.09.02.22279476

**Authors:** M Alvaro Berbís, David S. McClintock, Andrey Bychkov, Jerome Y Cheng, Brett Delahunt, Lars Egevad, Catarina Eloy, Alton B Farris, Filippo Fraggetta, Raimundo García del Moral, Douglas J. Hartman, Markus D Herrmann, Eva Hollemans, Kenneth A Iczkowski, Aly Karsan, Mark Kriegsmann, Jochen K Lennerz, Liron Pantanowitz, Mohamed E. Salama, John Sinard, Mark Tuthill, Jeroen Van der Laak, Bethany Williams, César Casado-Sánchez, Víctor Sánchez-Turrión, Antonio Luna, José Aneiros-Fernández, Jeanne Shen

## Abstract

**Background:** Artificial intelligence (AI) is rapidly fueling a fundamental transformation in the practice of pathology. However, AI’s clinical integration remains challenging, with no AI algorithms to date enjoying routine adoption within typical anatomic pathology (AP) laboratories. This survey gathered current expert perspectives and expectations regarding the role of AI in AP from those with first-hand computational pathology and AI experience.

**Methods:** Perspectives were solicited using the Delphi method from 24 subject matter experts between December 2020 and February 2021 regarding the anticipated role of AI in pathology by the year 2030. The study consisted of three consecutive rounds: 1) an open-ended, free response questionnaire generating a list of survey items; 2) a Likert-scale survey scored by experts and analyzed for consensus; and 3) a repeat survey of items not reaching consensus to obtain further expert consensus.

**Findings:** Consensus opinions were reached on 141 of 180 survey items (78.3%). Experts agreed that AI would be routinely and impactfully used within AP laboratory and pathologist clinical workflows by 2030. High consensus was reached on 100 items across nine categories encompassing the impact of AI on (1) pathology key performance indicators (KPIs) and (2) the pathology workforce and specific tasks performed by (3) pathologists and (4) AP lab technicians, as well as (5) specific AI applications and their likelihood of routine use by 2030, (6) AI’s role in integrated diagnostics, (7) pathology tasks likely to be fully automated using AI, and (8) regulatory/legal and (9) ethical aspects of AI integration in pathology.

**Interpretation:** This is the first systematic consensus study detailing the expected short/mid-term impact of AI on pathology practice. These findings provide timely and relevant information regarding future care delivery in pathology and raise key practical, ethical, and legal challenges that must be addressed prior to AI’s successful clinical implementation.

**Funding:** This research received no specific grant from any funding agency in the public, commercial, or not-for-profit sectors.

## INTRODUCTION

By virtue of its ability to “learn” from large volumes of electronic health record and image data without explicit programming, machine learning and artificial intelligence (AI) are set to fuel an unprecedented transformation in healthcare. In hospitals and clinics of the future, AI will become a cornerstone in the way care is delivered, contributing to more accurate diagnoses, increasingly agile, cost-effective, and standardized clinical workflows, and highly effective and personalized treatments.^1,2^ Excitement and expectations regarding the potential of AI to revolutionize healthcare have continued to build, as evidenced by the growing list of medical AI publications in the form of original research articles, review papers, health policy reports, white papers and consensus recommendations from professional societies, and coverage in the popular media.^1–16^ A recent survey of English-language articles indexed in PubMed showed a significant increase in the volume of medical AI research publications, from just 203 articles in 2005 to 12,563 in 2019.^3^

Pathology has attracted growing attention as an image-rich specialty likely to be strongly impacted by recent advances in AI. The development of machine learning-based tools for automated image analysis has led to a surge in AI applications promising to revolutionize current pathology workflows, as well as the advent of a new field, computational pathology.^7^ Between the years 2010-2020, approximately 23% (3,398) of all medical AI research publications were in pathology, making it the number one most-published specialty among the 17 specialties surveyed.^3^ A recent survey of the patent landscape from the years 1974–2021 yielded 523 patents relevant to the application of AI to digital pathology, with the primary application areas being whole-slide image (WSI) acquisition, segmentation, classification, and object detection.^17^ AI has been applied to several popular tasks in anatomic pathology (AP), including diagnosis, prognostication, and biomarker quantification, with key examples being automated assessment of prognostic biomarkers such as Ki-67 in breast cancer,^18–20^ automated tumor grading in prostate cancer,^21–25^ and diagnosis of metastatic breast cancer in lymph nodes.^26–29^ Other recently-explored AI applications in pathology have included tools for optimizing clinical laboratory workflows, such as automated quality control (QC).^30–32^

Much of the growth in computational pathology has been facilitated by the increased adoption of digital pathology, providing large amounts of WSI data as a prerequisite for practical AI model development and validation.^33^ Concurrently, use of digital workflows within pathology has provided practices with new platforms for testing and integrating computational pathology tools within AP workflows, generating greater interest in AI from pathologists and pathology trainees. A 2018 voluntary survey of 487 international pathologists and pathology trainees revealed a generally positive attitude towards AI, with nearly 75% expressing interest/excitement regarding the integration of AI tools into diagnostic pathology.^34^ Furthermore, 80% of respondents predicted the integration of AI-based assistance into AP workflows within the next 5–10 years.^34^ Numerous reviews have been published on the emerging and future applications of pathology AI to cancer diagnosis, prognostication, and treatment response prediction, metastasis detection in lymph nodes, single and multiplex biomarker quantification, tumor content/cellularity assessment for molecular testing, mutation status prediction, and a multitude of other tasks in pathology.^35–45^

Despite the recent progress and enthusiasm surrounding the application of AI to pathology, few algorithms are currently in routine clinical use,^37,46^ with a dearth of prospective multi-center, randomized trials present evaluating the impact of these algorithms in clinical settings.^47,48^ Further, ethical concerns have been raised regarding potential patient data privacy breaches, biased datasets producing systemic algorithmic bias, potential harm related to erroneous or misleading AI-generated outputs, and exacerbation of healthcare disparities due to unequal access to AI.^49^ All of these factors, along with hurdles related to regulatory approval and reimbursement for AI products, have contributed to a significant AI “translation gap” in pathology.^37^ While much has been written regarding the various opportunities and challenges surrounding AI in pathology, to date no systematic survey of opinions exists regarding the direct role of AI in pathology from the short to medium term perspective (the next decade) from those with dedicated expertise in digital and computational pathology. To address this knowledge gap, we conducted a consensus survey to gain detailed insight into the current challenges, expectations, and perspectives surrounding the role of AI in pathology, as seen from the point of view of an international panel of clinically active “early adopters” with hands-on experience developing and evaluating the clinical performance and utility of AI algorithms. For the survey, we chose to apply the Delphi method, a robust and widely accepted tool in medicine for building consensus among experts that has been shown to outperform standard statistical methods.^50,51^

A Delphi study involves a panel of experts who rate a series of statements, usually in a binary (e.g., agree/disagree) or semi-quantitative (e.g., Likert scale) manner. These statements might be produced after a systematic literature search or by surveying the same experts through a preliminary survey with open-ended questions (typically referred to as “Round 1”). Rating of statements is done over multiple consecutive rounds, during which the statements reaching the consensus criterion are omitted from subsequent rounds. Raters are then invited to reconsider their responses based on the group’s mean or median ratings from previous rounds. Studies are typically terminated when consensus has been reached, responses are stable, or after a pre-fixed number of rounds (usually two)^52^ to prevent expert fatigue. In contrast to typical single-round surveys, the Delphi technique relies on both the anonymization of responses to avoid bias and the use of experts and structured communication through an iterative pathway to reach more reliable conclusions.^53^

The goals of our study were to: 1) investigate the expected impact of AI on pathology; 2) forecast the extent of clinical AI implementation in the specialty by the end of this decade; and 3) provide specific insights into which technical, legal, regulatory, and ethical aspects of AI integration will require the most attention from pathology laboratories in the coming years. We expect the results of this study to be of broad interest to practicing pathologists, pathology trainees, pathology assistants and laboratory technicians, patients, physicians from other medical specialties, professional societies, hospital administrators, regulatory bodies, and researchers in academia, industry, and government.

## MATERIALS AND METHODS

### Expert Panel Recruitment

Candidates for the expert panel were selected by the research team using the following eligibility criteria: 1) professionals working within the specialty of pathology (anatomic and/or laboratory medicine) with an MD (or equivalent medical degree, such as an MBBS) and/or PhD, and 2) authorship of at least one peer-reviewed publication in the area of CPath/AI within the four years preceding the study (2016–2020) indexed in PubMed.

Invitations to participate in the study were sent in December 2020 to 39 candidates meeting the selection criteria. A total of 24 experts (60%) accepted and completed all three rounds of the survey. Detailed characteristics of the panelists are summarized in **table 1**. The majority of experts were practicing in North America (54%), with the remainder practicing in Europe, Japan, and New Zealand. The panelists were distributed across 10 different pathology subspecialties, with a third of panelists explicitly specializing in informatics, computational pathology, and/or digital pathology. Finally, the range of years in professional practice was mostly spread evenly across the panelists, with the majority having been in practice for 11–20 years (33%).

**Table 1.** Characteristics of the panelists

| Categories | n (%) |
| --- | --- |
| *Country of residence* |  |
| United States of America | 12 (50%) |
| The Netherlands | 2 (8·3%) |
| Spain | 2 (8·3%) |
| United Kingdom | 1 (4·2%) |
| Germany | 1 (4·2%) |
| Italy | 1 (4·2%) |
| Sweden | 1 (4·2%) |
| Portugal | 1 (4·2%) |
| Canada | 1 (4·2%) |
| Japan | 1 (4·2%) |
| New Zealand | 1 (4·2%) |
| *Area of subspecialization (multiple responses possible)* |  |
| Informatics, computational pathology, digital pathology | 8 (33·3%) |
| Hematopathology | 4 (16·7%) |
| Genitourinary pathology | 3 (12·5%) |
| Gastrointestinal pathology | 3 (12·5%) |
| Endocrine pathology | 2 (8·3%) |
| Dermatopathology | 2 (8·3%) |
| Surgical pathology | 2 (8·3%) |
| Molecular pathology | 2 (8·3%) |
| Thoracic pathology | 1 (4·2%) |
| Bone & soft tissue pathology | 1 (4·2%) |
| *Years of experience* |  |
| 0–10 | 7 (29·2%) |
| 11–20 | 8 (33·3%) |
| 21–30 | 7 (29·2%) |
| 31–40 | 2 (8·3%) |

### Delphi Study Development

To obtain a reliable consensus of opinions, the Delphi study was conducted over three rounds via a series of detailed questionnaires combined with controlled opinion feedback.^54^ The flowchart for the study is summarized in **figure 1**.

**Figure 1.**
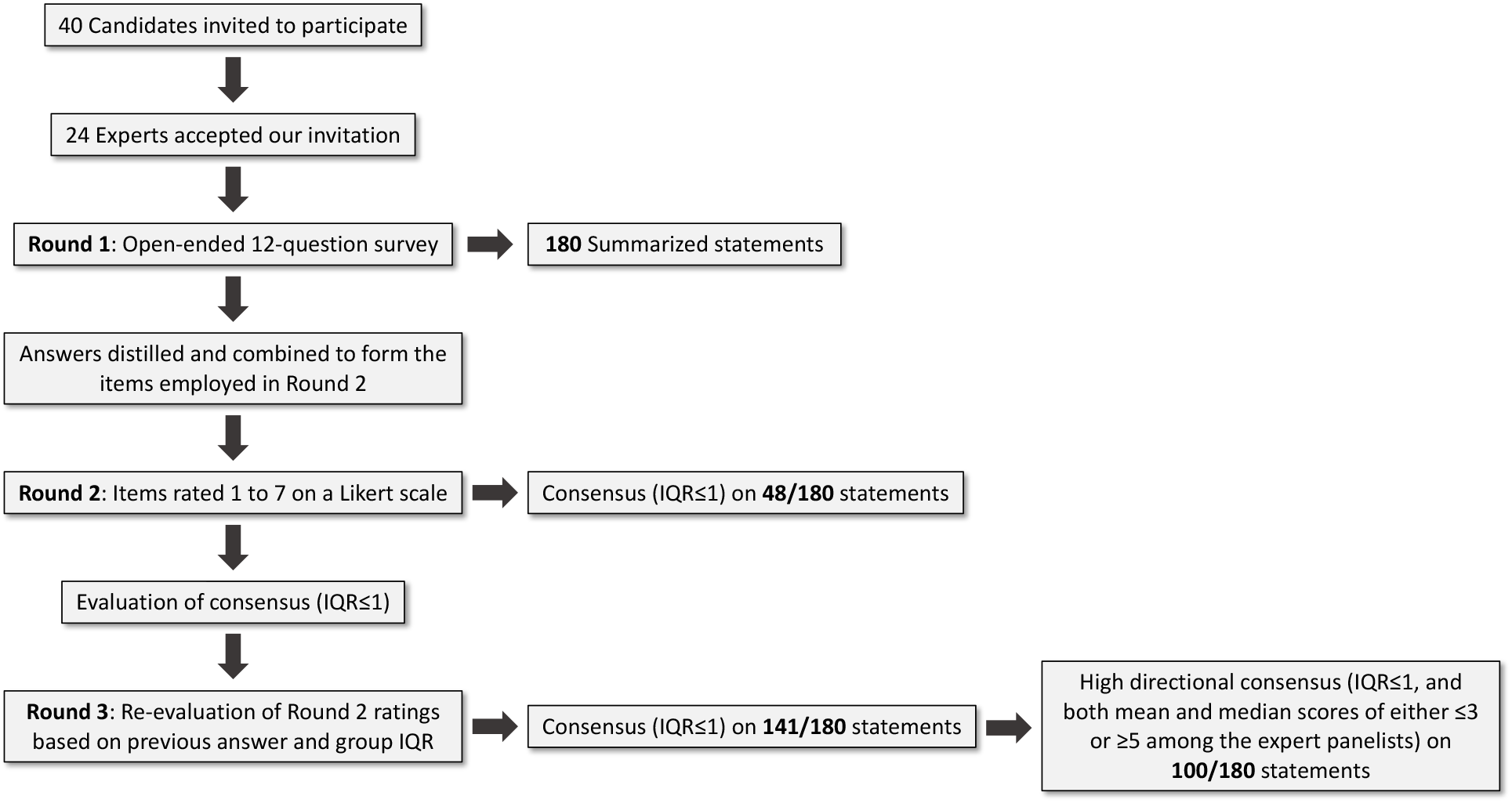
Flowchart illustrating the process of the present Delphi study

In Round 1, a preliminary review of the pathology AI literature, along with the research team’s empirical experience, was used to generate an open-ended questionnaire containing a set of 12 questions geared toward eliciting the ideas and opinions of the expert panelists regarding the following three topics: 1) forecasting the future of AI in pathology, 2) specific AI applications in pathology, and 3) ethical and regulatory aspects (**table 2**). The panelists’ responses to these open-ended questions were combined and distilled (all original answers were analyzed, and similar answers within the same category were grouped and summarized into a single statement, with the original wording used by the panelists reproduced wherever possible) into a series of questionnaire statements used in the subsequent survey rounds.

**Table 2.** Round 1 questions

| Section 1: Forecasts about the future (please answer according to what you believe will happen by 2030, instead of what you would like to see happen) |  |
| --- | --- |
| 1 | On what key performance indicators related to pathology do you believe AI will have a positive impact? |
| 2 | How do you think AI will impact the pathology workforce (jobs which will be created and jobs which will be destroyed) by 2030? |
| 3 | What new tasks will pathologists be involved in? |
| 4 | What new tasks will pathology technicians be involved in, or what existing tasks will they take on more responsibility for? |
| 5 | Which tasks currently performed by pathologists will be fully automated by AI by 2030? |
| **Section 2: Applications of AI in pathology** (please cite any existing or potential AI-based tools which, in your opinion, would bring value to pathologists. Be as specific as possible) |  |
| 6 | In what ways can AI be used to improve diagnostic precision? |
| 7 | In what ways can AI be used to speed up or facilitate the work of pathologists? |
| 8 | What examples of AI tools or applications would bring value to the analysis and interpretation of histological images? |
| 9 | What examples of AI tools or applications would bring value to other aspects of the laboratory workflow? |
| 10 | In what ways can AI be used to bring value to integrated diagnostics? (integrated diagnostics refers to the convergence of two or more diagnostic techniques, such as pathology, radiology, genomics) |
| **Section 3: Ethical and regulatory aspects** |  |
| 11 | What regulatory challenges will have to be overcome for the generalized adoption of AI in the pathology setting? |
| 12 | What ethical issues could arise from the use (and potential misuse) of AI in the pathology setting? |

AI, artificial intelligence.

In Round 2, the panelists were asked to rate each of the questionnaire statements on a 7-point Likert scale (from 1 to 7), with different scores designed to fit different categories of questions. For example, a score of 1 might indicate “Very strongly disagree”, “Impossible” (with regard to likelihood of an event occurring), “Disappear”(with regard to job number or availability), or “Not involved at all” (with regard to degree of involvement in specific tasks), whereas a score of 7 might indicate “Very strongly agree”, Certain”, “Dramatically increase”, or “Daily involvement”, depending on the question, with higher scores generally representing a more favorable opinion toward the future role or impact of AI on Pathology (**table 3**).

**Table 3.** Likert scales for the different sections (7-point)

| Point Score | Agreement scale | Probability scale | Job number variation scale | Involvement scale |
| --- | --- | --- | --- | --- |
| 1 | Very strongly disagree | Impossible | Disappear | Not involved at all |
| 2 | Strongly disagree | Very unlikely | Greatly decrease | Rarely involved |
| 3 | Disagree | Unlikely | Somewhat decrease | Somewhat involved |
| 4 | Neither agree nor disagree | Even chance / neutral | Remain the same | Sometimes involved |
| 5 | Agree | Likely | Somewhat increase | Often involved |
| 6 | Strongly agree | Very likely | Greatly increase | Routinely involved |
| 7 | Very strongly agree | Certain | Dramatically increase | Daily involved |

In Round 3, the panelists were asked to re-rate all statements which did not reach consensus during Round 2. Consensus on statements was defined, a priori, as having an interquartile range (IQR) of less than 1 (IQR ≤ 1) for ratings along the 7-point Likert scale.^55,56^ During Round 3, the panelists were shown their Round 2 ratings on each statement, along with the median and IQR for the group as a whole, and finally given the option to change their previous ratings, if desired.

All questionnaires were completed via an online Google Form (Google Inc, Mountain View, CA, USA), with individualized links to the form e-mailed to each panelist. The survey participants remained anonymous to one another during all three Rounds, with each participant able to view only their own responses to each statement during Rounds 1 and 2, along with the anonymized group medians and IQRs during Round 3.

### Statistical Analysis

To examine whether any significant differences in panelist scores were present based on gender, practice location, pathology subspecialty, or years in practice, Wilcoxon rank-sum tests (two-tailed, alpha = 0.05 significance criterion) were performed. All analyses were performed using Microsoft Excel (Microsoft Corporation, Redmond, WA, USA) and STATA v16 (StataCorp LLC, College Station, TX, USA).

## RESULTS

### Survey Rounds

The unstructured first Delphi round allowed the panelists relative freedom in expressing their individual thoughts on topics they felt were important or relevant to the future of AI in pathology over the next decade. This resulted in the generation of 180 summative statements spanning the following nine categories: 1) impact of AI on key performance indicators (KPIs), 2) impact of AI on the pathology workforce, 3) impact of AI on pathologist tasks, 4) impact of AI on pathology technician tasks, 5) specific applications of AI in pathology, 6) role of pathology AI in integrated diagnostics, 7) pathology tasks likely to be fully automated by AI, 8) regulatory and legal aspects of AI integration, and 9) ethical aspects of AI integration.

One of the motivations behind using the Delphi method was its ability to achieve greater consensus (a reduction in variance across rounds, as measured by the IQR) among a limited number of expert panelists. After Round 2, responses to 48 of the 180 (26.7%) statements reached consensus (IQR ≤ 1). After Round 3, the panelists had reached consensus on 141 of the 180 (78.3%) statements. The mean and median Likert scores across the panelists for each of the 180 statements ranged from 3.04 to 6.83 (mean), and from 3 to 7 (median), respectively. Tables 4 through 10 present the 100 statements (spread across seven general categories) that achieved high directional consensus (defined as IQR ≤ 1, *and* both mean and median scores of either ≤ 3 or ≥ 5) among the expert panelists. For these 100 statements, two-tailed Wilcoxon rank sum tests demonstrated no significant differences in Likert scores between the various panelist comparison groups (female vs. male, North American vs. non-North American, subspecialty informatics or computational/digital pathology vs. other subspecialty, and ≤ 10 years vs. ≥ 11 years in practice) on 85 of the 100 statements. The remaining 15 statements are further discussed in the corresponding sections below, which summarize the most significant results of the survey according to topic/category

**Table 4.**
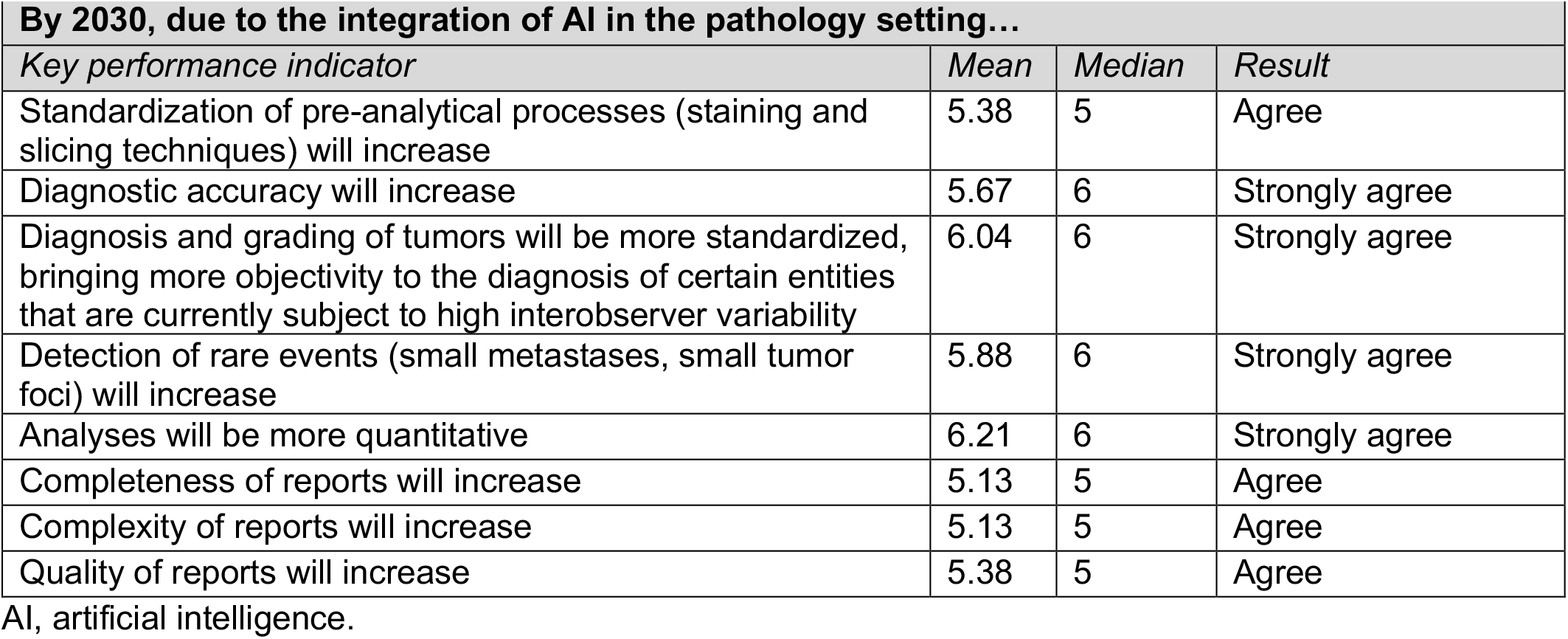
Statements on which high directional consensus was reached regarding the impact of AI on pathology key performance indicators (KPIs)

| By 2030, due to the integration of AI in the pathology setting... |  |  |  |
| --- | --- | --- | --- |
| Key performance indicator | Mean | Median | Result |
| Standardization of pre-analytical processes (staining and slicing techniques) will increase | 5.38 | 5 | Agree |
| Diagnostic accuracy will increase | 5.67 | 6 | Strongly agree |
| Diagnosis and grading of tumors will be more standardized, bringing more objectivity to the diagnosis of certain entities that are currently subject to high interobserver variability | 6.04 | 6 | Strongly agree |
| Detection of rare events (small metastases, small tumor foci) will increase | 5.88 | 6 | Strongly agree |
| Analyses will be more quantitative | 6.21 | 6 | Strongly agree |
| Completeness of reports will increase | 5.13 | 5 | Agree |
| Complexity of reports will increase | 5.13 | 5 | Agree |
| Quality of reports will increase | 5.38 | 5 | Agree |

AI, artificial intelligence.

### Impact of AI on Pathology Key Performance Indicators (KPIs)

In general, there was agreement that AI would help improve laboratory KPIs, such as turnaround time, diagnostic accuracy, the detection rate for rare events (for example, small foci of primary or metastatic tumor), and quality of diagnostic reports (**table 4**). However, it was also anticipated that histopathologic analyses would become more quantitative, and that diagnostic reports would become more complex. There was also agreement that AI would lead to greater standardization of diagnostic and pre-analytical processes, such as tissue sectioning, staining techniques, and workflows. As a result of these anticipated improvements in KPIs, it was expected that there would be an increase in the satisfaction of referring physicians. A statement regarding the likelihood of the cost per case decreasing with the use of AI failed to reach consensus, although a majority of the experts predicted the cost per case would not decrease with medium term AI use (within the next 8-10 years). In addition, the panelists did not reach consensus on whether the number of second-opinion consultations would decrease as a result of AI adoption.

**Table 4** presents those statements regarding the impact of AI on pathology KPIs for which there was high directional consensus among the panelists. Overall, these results projected, by the year 2030, growth in computational pathology as a subspecialty with AI applications assisting pathologists in making diagnoses that are more accurate, standardized, objective, quantitative, and complete.

### Impact of AI on the Pathology Workforce and Specific Tasks

There was agreement that the number of jobs for pathologists, as well as administrative staff, would likely remain the same. However, a modest increase in the number of jobs for technicians and information technology (IT) professionals was expected. While the size of the overall pathology job market was not expected to vary greatly due to the adoption of AI, the types and frequencies of tasks performed by pathologists and pathology laboratory technicians were expected to change significantly. There was agreement that AI would facilitate subspecialization in pathology, with the number of pathologists specializing in computational pathology greatly increasing. Tables 5 and 6 present the expected impact of AI on the pathologist (**table 5**) and pathology laboratory technician (**table 6**) workforces and associated tasks.

**Table 5.** Statements on which high directional consensus was reached regarding the impact of AI on pathologist workforce and tasks

| By 2030, due to the integration of AI in the pathology setting... |  |  |  |
| --- | --- | --- | --- |
| Task | Mean | Median | Involvement/Agreement Level |
| The number of jobs for IT staff will... | 5.54 | 5 | Somewhat increase |
| The number of specialized “computational” pathologists will greatly increase | 5.75 | 6 | Strongly agree |
| Pathologists will be more involved in diagnostic tumor boards | 5.58 | 6 | Strongly agree |
| Pathologists will be more involved in multidisciplinary conferences | 5.63 | 6 | Strongly agree |
| Pathologists will be more involved in research activities | 5.42 | 5 | Agree |
| Pathologists will be spending more time in the study of rare lesions | 5.13 | 5 | Agree |
| By 2030, due to the integration of AI in the pathology setting, the degree of involvement of pathologists in these tasks will be... |  |  |  |
| Digital pathologic diagnosis without the use of physical glass slides | 5.58 | 6 | Routine |
| Interpretation of computationally derived measurements and evaluations | 6.08 | 6 | Routine |
| Collaboration with EHR teams regarding the use of laboratory data for a wide range of clinical decision support tools | 5.25 | 5.5 | Routine |
| Evaluating different kinds of AI software and deciding whether these are appropriate for their workflow | 5.54 | 6 | Routine |
| Validation and QA/QC of AI solutions | 5.63 | 6 | Routine |
| Validation and QA/QC of AI-rendered diagnoses | 5.88 | 6 | Routine |
| Defining new categories of patients, based on new data made available through AI | 5.04 | 5 | Often |

AI, artificial intelligence; IT, information technology; EHR, electronic health record; QA/QC, quality assurance/quality control.

**Table 6.** Statements on which high directional consensus was reached regarding the impact of AI on pathology lab technician workforce and tasks

| By 2030, due to the integration of AI in the pathology setting, the degree of involvement of pathology laboratory technicians in these tasks will be... |  |  |  |
| --- | --- | --- | --- |
| Task | Mean | Median | Involvement Level |
| Operation of digital slide scanners, digitization, and image management | 6.25 | 7 | Daily |
| QA/QC of digitized images | 6.08 | 6.5 | Daily |
| Digital pathology support for pathologists and other users, such as device calibration | 5.88 | 6 | Routine |
| Assessing histology consistency, i.e., re-addressing SOPs to make slides and corresponding images more suitable for AI (more consistent tissue and staining quality) | 5.83 | 6 | Routine |
| Validation and QA/QC of AI-rendered diagnoses | 5.17 | 5 | Often |

AI, artificial intelligence; SOP, standard operating procedure; QA/QC, quality assurance/quality control.

There was agreement that, by 2030, pathologists would be routinely involved in new tasks related to the incorporation of AI into their workflows, such as interpreting diagnostic outputs from AI algorithms, evaluating AI software, and performing validation and quality assurance/quality control (QA/QC) for AI solutions and AI-rendered diagnoses. There was also agreement pathologists would be directly participating in the design and development of AI solutions and, based on the availability of new data generated by AI, they would also be contributing to the definition of new categories of patients. As a result of AI adoption, pathologists were also expected to be more involved in ancillary activities, such as research and participation in multidisciplinary conferences and tumor boards. The panelists also agreed pathologists would sometimes be more involved in meeting directly with patients, including more frequent inclusion in patient treatment decision-making.

Compared to the panelists who had been in practice for 11 years or longer, those in practice for 10 or fewer years (7 panelists) more strongly agreed that digital pathologic diagnosis without the use of physical glass slides would be routine by 2030 (*p* = 0.041), with a median score of 7 (compared with a median score of 6, for those with 11 or more years of practice) and mean score of 6.43 (compared to a mean score of 5.23, for those with 11 or more years of practice). The same group also more strongly agreed that interpretation of computationally derived measurements and evaluations would be routine (*p* = 0.048), with a median score of 7 (compared with a median score of 6, for those with 11 or more years of practice) and mean score of 6.71 (compared to a mean score of 5.82, for those with 11 or more years of practice).

The work of pathology technicians was also expected to undergo major changes as a result of AI adoption (**table 6**). Technicians would be routinely involved in tasks related to new digital and computational workflows, such as scanner operation, device calibration, and QA/QC of digitized images. There was no consensus, however, as to whether technicians would be directly participating in AI-assisted diagnosis, although a slight majority of the panelists surveyed thought this would be the case.

The panelists subspecializing in informatics or computational/digital pathology (8 panelists) more strongly felt that pathology laboratory technicians would routinely be providing digital pathology support for pathologists and other users (for example, by performing device calibration and other tasks) (*p* = 0.005), with a median score of 7 and a mean score of 6.63 (as compared to a median of 6 and a mean of 5.50 for those not subspecializing in informatics or computational/digital pathology). Similarly, this group also felt more strongly that pathology laboratory technicians would be routinely involved in assessing and improving the consistency of histologic tissue preparation and staining to make images more suitable for AI (*p* = 0.049), with a mean score of 6.25 (as compared to a mean of 5.63, with the same median of 6, for those not subspecializing in informatics or computational/digital pathology).

### Applications of AI to Pathology and Integrated Diagnostics

AI was expected to positively impact many aspects of the pathology workflow. The panelists proposed several specific applications they thought would likely be in routine use in pathology laboratories by the year 2030 (**table 7**). Regarding the analysis and interpretation of histologic images, routine applications included algorithms for the identification of hotspot areas (for example, during mitotic counts), detection of microorganisms (such as acid-fast bacilli and *Helicobacter pylori*) and cancer, and assistance with tumor grading. The panelists were also certain AI would be in routine use for the automated quantification of immunohistochemical (IHC) and immunofluorescent (IF) biomarker stains, such as Ki-67, estrogen receptor (ER), progesterone receptor (PgR), and programmed cell death ligand 1 (PD-L1), as well as the counting of mitotic figures and lymphocytes on hematoxylin and eosin (H&E) stained tissue, and the identification of lymph node metastases and, in particular, micrometastases.

**Table 7.**
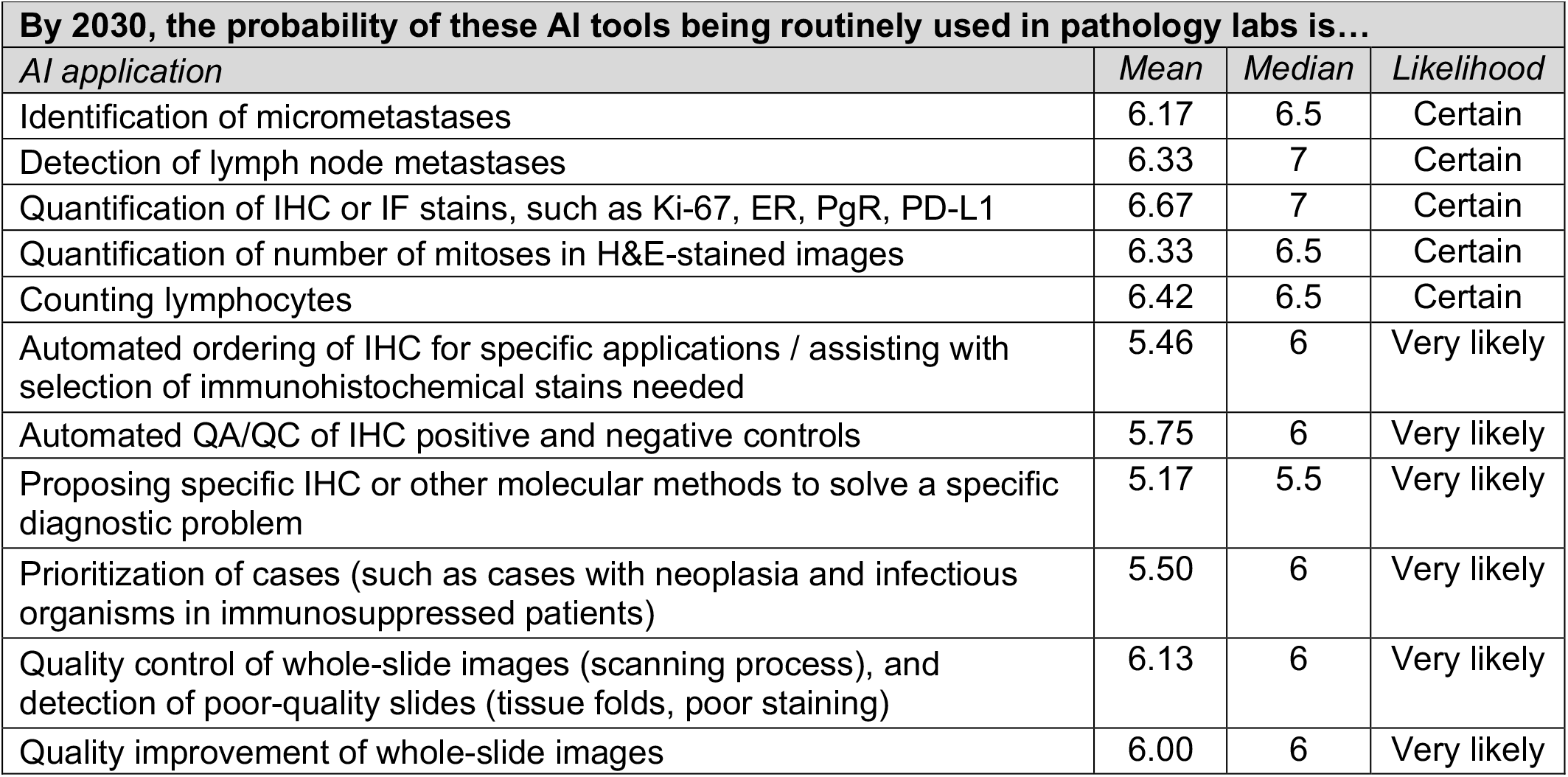
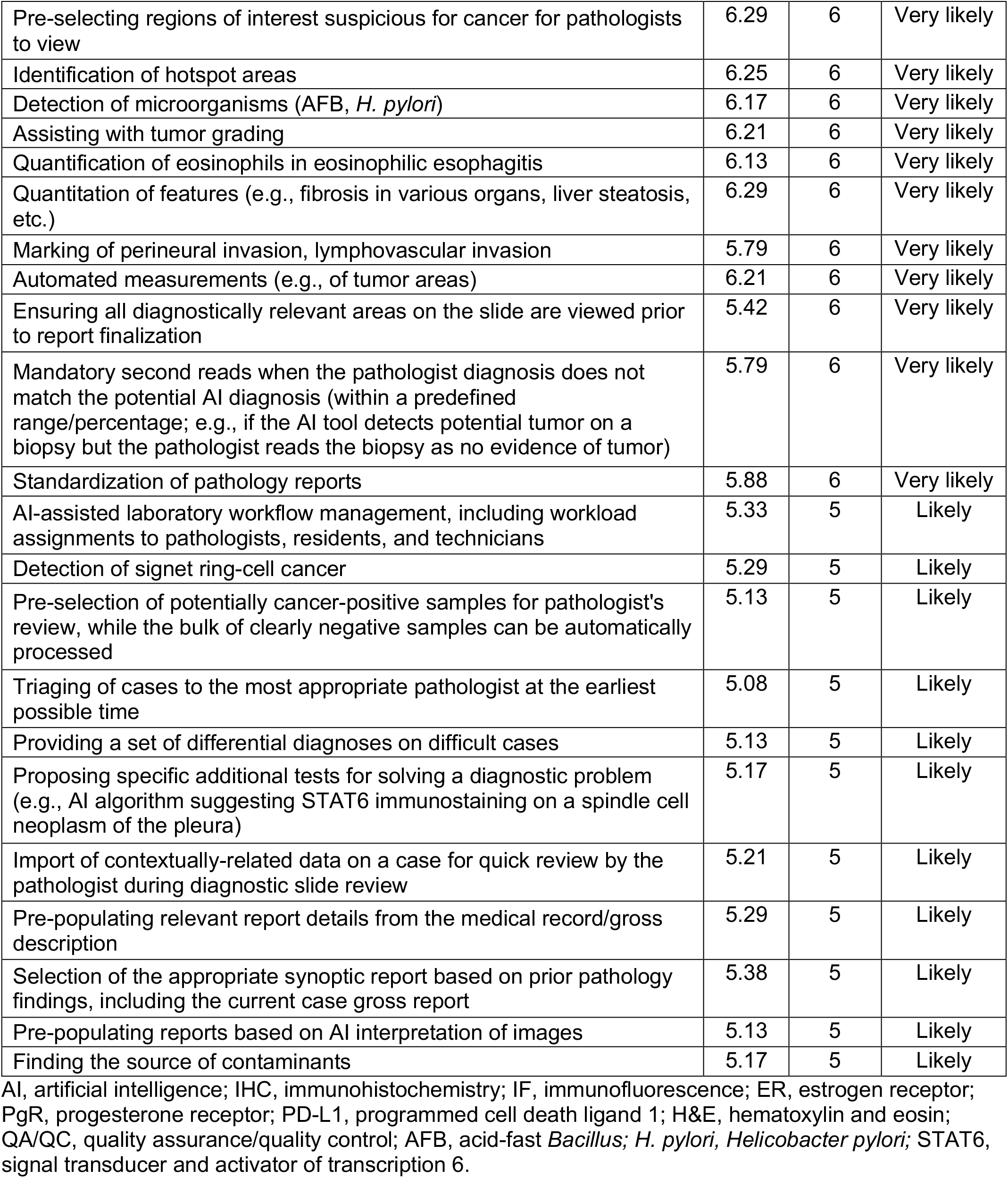
Statements on which high directional consensus was reached regarding AI applications in pathology

Tasks which are currently performed manually, but which were expected to be replaced by AI automation by the year 2030, included the provision of size measurements (e.g., tumors) and the detection of perineural and lymphovascular invasion in malignancies. In addition, it was expected that AI-based computational/virtual staining would replace the need for multiplex histochemistry and IHC/IF.

Within a decade, AI was expected to increase diagnostic efficiency by prioritizing regions of interest suspicious for cancer involvement for pathologists to review first, pre-populating relevant diagnostic report fields using data extracted from the medical record and/or gross description, and pre-populating diagnostic reports based on AI interpretation of whole-slide images. In addition, AI was expected to assist pathologists with making more accurate diagnoses by importing contextually relevant clinical history and other data related to a case for quick review by the pathologist during case sign-out, providing a set of differential diagnoses, and prompting second reads on cases where there was a discrepancy between the pathologist diagnosis and the diagnosis rendered by the AI algorithm.

The panel also foresaw a significant impact of AI on other aspects of the laboratory workflow. Routine AI-assisted prioritization of cases for pathologist review, suggestions for ancillary stains to be performed, and automated ordering of additional stains were thought to be very likely by 2030 (**table 7**). In terms of differences in opinion between the various panelist subgroups, those subspecializing in informatics or computational/digital pathology less strongly believed that AI would routinely be used for the quantification of eosinophils in eosinophilic esophagitis (*p* = 0.049), with a mean score of 5.75, as compared to a mean of 6.31 (with the same median of 6), for the other subspecialists.

AI was expected to foster the integration of pathology with other diagnostic modalities, such as radiology and genomics, by: 1) identifying histologic regions of interest to be sampled for genomic testing; 2) correlating morphologic and genomic information to help interpret genetic aberration; 3) comparing tumor extent on histology slides and radiologic images; and 4) interpreting tumor treatment response in radiologic images, based on pathology data. Multimodal-AI was also expected to enable the combination of diverse types of diagnostic data (gross/macroscopic, microscopic, radiologic, and genomic) in a single interface, and to facilitate the use of integrated diagnostic reports for certain diseases, such as prostate cancer (**table 8**). Consequently, it was felt AI-powered integrated diagnostics would likely lead to significant advances in the personalization of healthcare by creating new categories of patients based on differential risk stratification (prognostic) roadmaps and prediction of clinical outcomes.

**Table 8.**
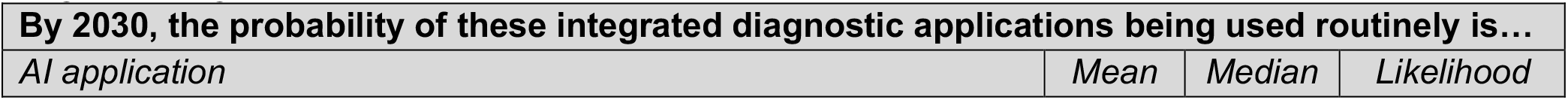
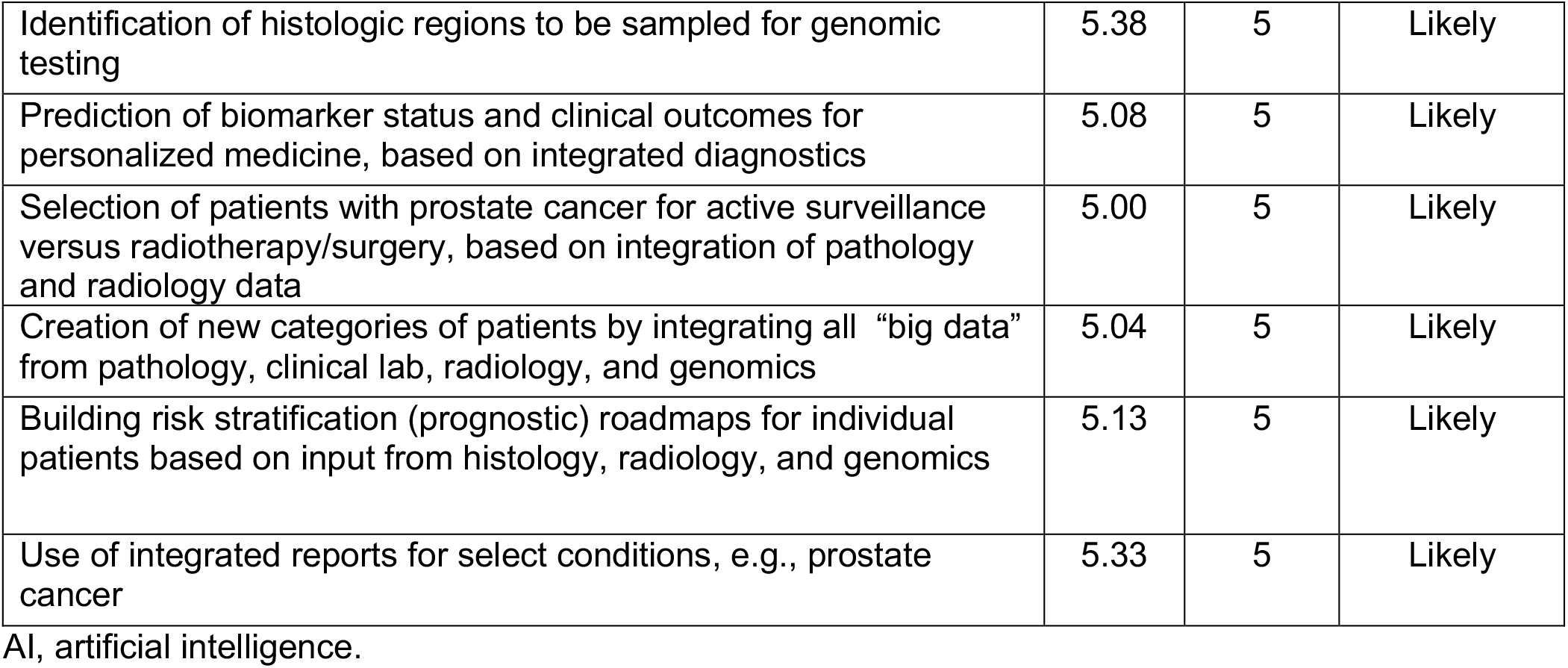
Statements on which high directional consensus was reached regarding the role of AI in integrated diagnostics

The panelists with greater practice experience (≥ 11 years) more strongly believed the integration of pathology and radiology data would routinely be used to select patients for active surveillance versus radiotherapy/surgery in prostate cancer (*p* = 0.044; equal medians of 5 but mean of 5.29 versus 4.29 for those with ≥11 years and ≤ 10 years of practice experience, respectively). The panelists with more practice experience more strongly believed that AI would routinely be used to build risk stratification (prognostic) roadmaps for patients based on histologic, radiologic, and genomic input data (*p* = 0.020; equal medians of 5, means of 5.41 versus 4.43 for those with ≥ 11 years and ≤ 10 years of practice experience, respectively).

The panelists felt it was likely that, by 2030, AI would not simply assist with, but would fully replace, pathologists on several manually-performed tasks. Among these were stain selection; measurement tasks; prioritization and triage of cases; screening for microorganisms; colorectal polyp, cervical cytology, and lymph node screening; and grading of breast and colorectal cancers (**table 9**). Assigning work to pathologists, trainees, and technicians, as well as triaging cases to the appropriate pathologist, were also deemed likely to be fully automated by AI.

**Table 9.** Statements on which high directional consensus was reached regarding pathology tasks expected to be fully automated by 2030

| By 2030, the probability of these tasks being fully delegated to AI in pathology labs is... |  |  |  |
| --- | --- | --- | --- |
| Task | Mean | Median | Likelihood |
| Verification of positive and negative controls for IHC | 5.71 | 6 | Very likely |
| Prioritization of cases | 5.54 | 6 | Very likely |
| Triage of cases to appropriate pathologists | 5.46 | 6 | Very likely |
| Contextual data lookup on patients from the EHR relevant to the pathology case being reviewed | 5.25 | 6 | Very likely |
| Slide QC (e.g., detection of tissue folds and tears, stain quality evaluation, etc.) | 5.88 | 6 | Very likely |
| Screening for microorganisms, such as AFB and *H. pylori* | 5.96 | 6 | Very likely |
| Screening of colorectal polyps | 5.58 | 6 | Very likely |
| Cervical cytology screening | 6.21 | 6 | Very likely |
| Screening lymph nodes for metastases | 5.83 | 6 | Very likely |
| Measurement tasks | 6.17 | 6 | Very likely |
| Quantification of IHC or IF stains, such as Ki-67, ER, PgR, PD-L1 | 6.29 | 6 | Very likely |
| Quantification of mitotic count on H&E-stained images | 6.08 | 6 | Very likely |
| Bone marrow differential counts | 5.54 | 6 | Very likely |
| MIB-1 scoring | 6.04 | 6 | Very likely |
| Assessing extent of liver steatosis and fibrosis | 5.54 | 6 | Very likely |
| Screening of tissues with a cancer diagnosis to select regions for tissue coring or macroscopic dissection | 5.08 | 5 | Likely |
| Slide screening for regions of interest | 5.13 | 5 | Likely |
| Grading of breast cancer | 5.42 | 5 | Likely |
| Grading of colorectal cancer | 5.33 | 5 | Likely |

AI, artificial intelligence; IHC, immunohistochemistry; EHR, electronic health record; QC, quality control; AFB, acid-fast *Bacillus*; *H. pylori*, *Helicobacter pylori*; IF, immunofluorescence; ER, estrogen receptor; PgR, progesterone receptor; PD-L1, programmed cell death ligand 1; H&E, hematoxylin and eosin.

The panelists practicing outside of North America (11 panelists) thought it more likely that colorectal polyp screening would be fully delegated to AI by the year 2030 (*p* = 0.047; mean = 6 and median = 6 for non-North American panelists, versus mean = 5.23 and median = 5 for North American panelists). Those with more practice experience (≥ 11 years) thought it more likely that mitotic counts on H&E-stained images would be fully delegated to AI (*p* = 0.036; equal medians of 6 but mean of 6.29 versus 5.57 for those with ≥ 11 years and ≤ 10 years of practice experience, respectively).

### Regulatory and Ethical Aspects of AI Integration in Pathology

The panelists foresaw significant regulatory and ethical challenges posed by the integration of AI in pathology (**table 10**). While some panelists initially suggested such issues might not pose a challenge to the application of AI in pathology, since the pathologist (or another physician) always takes the ultimate responsibility for the diagnostic, therapeutic or prognostic use of AI, the final consensus opinion overruled that initial suggestion. In principle, the panel agreed that both primary diagnostic and secondary (e.g., advisory/assistive) algorithms would have to meet strict regulatory requirements.

**Table 10.**
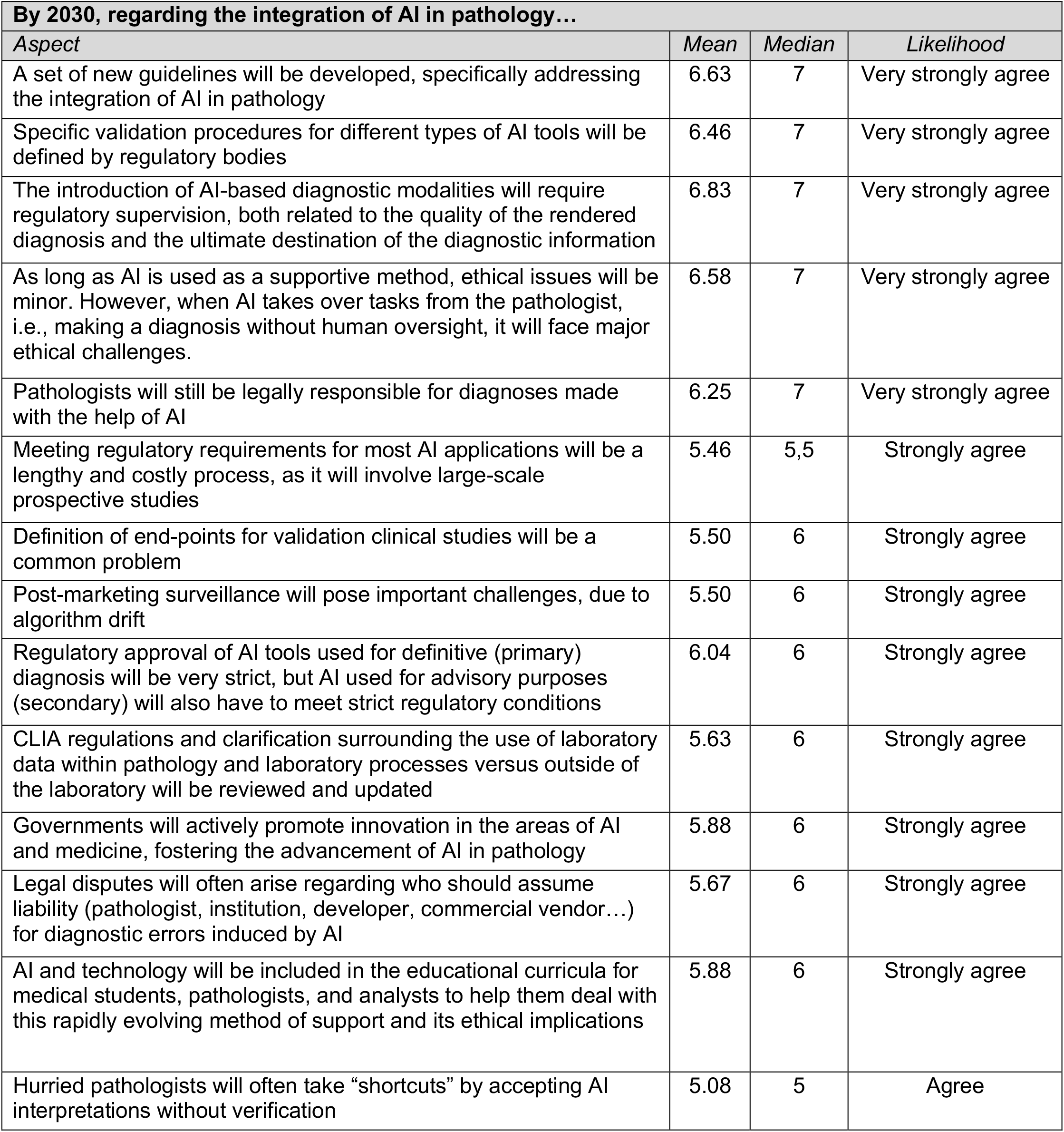
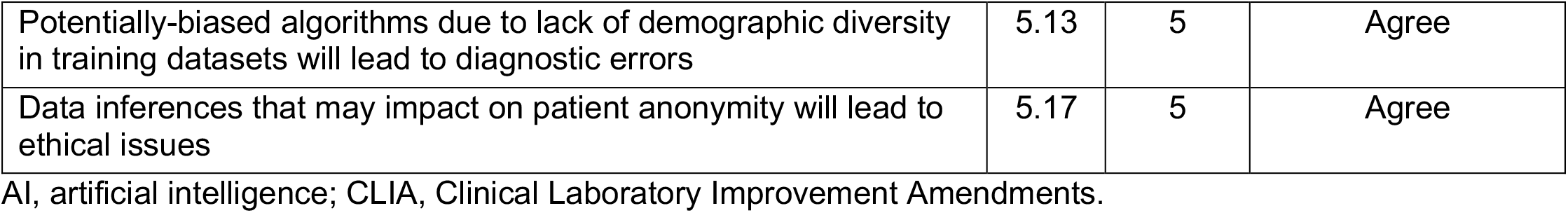
Statements on which high directional consensus was reached regarding regulatory and ethical aspects

There was agreement that regulatory bodies would create new guidelines addressing the integration of AI into pathology, which would provide specific validation procedures and simplify regulatory pathways for AI tools, although regulatory clearance of AI software would still be a lengthy and costly process.

There was also agreement that the regulatory approval of adaptive algorithms, which continuously evolve in response to new input data, would be possible, but that algorithm drift would pose important challenges that would need to be tackled by close post-market surveillance. It was also anticipated that legal disputes would arise regarding who should assume liability for diagnostic errors induced by AI, with pathologists still being held legally responsible for AI-assisted diagnosis.

With regard to differences in opinion between the various panelist subgroups, the North American panelists more strongly believed that CLIA regulations and clarification surrounding the use of laboratory data within pathology, as well as laboratory processes, would need to be reviewed and updated (*p* = 0.031; median = 6 vs. median = 5 and mean = 6 vs. mean = 5.18 for North American vs. other panelists, respectively). Those subspecialized in informatics or computational/digital pathology less strongly believed that legal disputes would often arise regarding who should assume liability for diagnostic errors induced by AI (*p* = 0.018; mean = 4.88 and median = 5), compared to those not subspecialized in those areas (mean = 6.06 and median = 6).

Overall, the panel acknowledged there would be major ethical issues arising from the full delegation of tasks to AI algorithms, believing it likely that hurried pathologists would often accept AI interpretations without sufficient verification. On the other hand, there was disagreement between the panelists regarding whether the black box nature of AI algorithms would cause pathologists to often make diagnoses without enough clinical explainability. The panelists who had been in practice fewer years (≤ 10 years) more strongly believed that ethical issues would result from data inferences which might compromise patient anonymity (*p* = 0.004; mean = 6 and median = 6), compared to those in practice ≥ 11 years (mean = 4.82 and median = 5).

Finally, there was general consensus that ethical issues were expected to arise due to: 1) risk for diagnostic error resulting from the use of potentially biased algorithms trained on datasets lacking sufficient demographic diversity; and 2) the lack of proper informed consent when using patient data (which the panel agreed would become a common practice). Nevertheless, the panelists expected official regulatory bodies would do their part to overcome the aforementioned ethical and legal challenges, and funding bodies would actively promote innovation in AI and medicine, thereby fostering the advancement of AI in pathology. It was also anticipated AI would be integrated into both medical school and continuing medical education curricula for pathologists in current practice, in order to help them adapt to this rapidly evolving area and its associated legal and ethical implications.

## DISCUSSION

From this first ever consensus study of a panel of 24 international experts with first-hand experience working in the field of computational pathology and AI (almost all of whom are also pathologists in active clinical practice), we obtained specific insight into consistently agreed-upon opportunities and challenges, as well as perspectives and predictions, relevant to the expected role of AI in pathology over the next decade. Our panel was composed of experts of both genders, practicing in 11 different countries, specializing in a wide range of pathology subspecialties, and with a wide range of years in practice (with all panelists having attending pathologist/faculty status). Despite this diversity in the expert panel, the panelists were able to reach consensus agreement on 140 (78.3%) of the 180 items surveyed.

There was particularly strong consensus AI would improve the KPI of diagnostic accuracy, at least partially by assisting with the detection of rare events (such as small tumor foci and metastases), standardizing the diagnosis and grading of tumors, and making histopathologic analyses more quantitative. There was also particularly strong consensus that the number of specialized computational pathologists would greatly increase, and that pathologists would be more involved in diagnostic tumor boards and multidisciplinary conferences as a result of AI integration. Significant changes in the types of tasks routinely performed by a pathology laboratory technician were also expected.It was projected that laboratory technicians would be operating digital slide scanners and performing digital image management and QA/QC daily.

It was felt to be *almost certain* that specific pathology AI applications would be in routine use by 2030 – namely, algorithms for the identification of lymph node macro- and micro-metastases, IHC/IF stain quantification (including Ki-67, ER, PgR, and PD-L1), mitosis quantification, and lymphocyte quantification. It was also felt to be *very likely* that algorithms would be in routine use for: 1) specific pre-analytical tasks, such as automated QA/QC of histologic preparation, WSI quality, and IHC controls, automated suggestion or ordering of ancillary stains and molecular tests, and automated case prioritization; 2) specific analytical tasks, such as pre-selection of cancer regions of interest and hotspot areas, microorganism detection, tumor grading, eosinophil and feature quantification, tumor area measurement, identification of lymphovascular and perineural invasion, diagnostic report standardization, and ensuring that all diagnostically-relevant areas in a WSI are viewed prior to report finalization; and (3) specific post-analytical tasks, such as enforcement of mandatory second reads when there is a significant discrepancy between the pathologist-rendered and AI-rendered diagnoses. The panelists felt it *very likely* that many of the preceding tasks, along with colorectal polyp and cervical cytology screening, case triage and assignment to pathologists, and contextual electronic health/medical records data lookup, would no longer be performed by pathologists in the year 2030, as they will have been *fully delegated* to AI. These predictions are consistent with the relatively high representation of many available applications in the existing pathology AI research literature. ^29,31,41,43,57–63^

At the same time, it is interesting to note that many of the applications projected to be in routine use within the next decade address basic or mundane tasks already being performed by pathologists, rather than advanced or “aspirational” tasks pathologists do not currently perform (such as prediction of molecular biomarker status or clinical outcomes directly from morphologic features on H&E-stained slides). The relative projected likelihoods of these two dominant categories of AI applications (“basic” versus “aspirational”) being in routine use by 2030 seems, in many ways, contrary to the relative degree of attention paid to these categories by the research community and industry stakeholders. ^35,41,64–66^ For example, in a recent survey from 2021,^67^ 75 computational pathology domain experts from academia and industry (48% with medical and 52% with non-medical backgrounds) were asked to provide their subjective rankings regarding the degree of interest, importance, and/or promise of 12 solid tumor-specific pathology AI applications. Among the applications surveyed, those falling within the advanced/aspirational categories were consistently rated the most highly, while those falling within the more basic/mundane categories (such as QA/QC, tumor grading, and subtyping) were rated the lowest, with the most highly-rated application being prediction of treatment response directly from H&E images, followed by prediction of genetic mutations, gene expression, and survival directly from H&E images, respectively. Although “degree of interest/importance/promise” is not exactly equivalent to the “likelihood of routine near-term adoption”, the somewhat discrepant findings between the two surveys, each of which targeted different participant populations, suggests that those with non-medical and/or industry backgrounds (who collectively formed the majority of participants in the cited study) might be more optimistic about the near-term role and importance of advanced/”aspirational” AI applications in pathology than pathologists with first-hand experience working in the areas of computational pathology/AI, who tend to have a more practical perspective regarding those AI applications likely to enjoy widespread adoption in the near to medium term. It was unclear from our study why more advanced/”aspirational” AI tools were not predicted to be in routine use within the next decade.

Perhaps, it was felt that such tools, though interesting from a research standpoint, would never reach the level of performance required for clinical adoption. Or, in order to have a sufficient level of performance for these tools, the fundamental AI infrastructure addressing the “basic” tasks would have to be completed first, paving the way for full diagnostic AI algorithm clinical validation and regulatory clearance. It remains for future studies to explore the reasons underpinning the results observed in this (and other) surveys regarding clinical AI.

Only a few other studies have sought to survey the opinions of various stakeholders regarding AI in pathology. In a 2018 survey of 363 practicing pathologists and 124 pathology trainees from 54 countries (46% female, 56% practicing in the United States, Canada, or the United Kingdom, and 43% in practice for ≤10 years) on general perspectives regarding the integration of AI into diagnostic pathology, 81% of respondents predicted that AI would be integrated into pathology workflows within the next 5–10 years.^34^ In the same survey, 38% of respondents felt that AI would have no impact on pathologist employability, 42% felt that it would create new positions and improve employment prospects, and 20% were either concerned or extremely concerned that AI would displace them from their jobs. Approximately 72% of the respondents also felt that AI tools could increase or dramatically increase diagnostic efficiency, with the remaining 28% being unsure of AI’s impact on efficiency, or of the opinion that AI would have either no impact or a negative impact on efficiency. In contrast to the more heterogeneous respondent demographic in that survey, the participants in our survey were composed almost exclusively of practicing pathologists with experience in computational pathology/AI. Although the consensus perspectives from our survey were more optimistic regarding the impact of AI on the pathologist workforce, there was similar reservation regarding whether AI integration would truly lead to increased efficiency, in terms of cost per case (one of the KPI’s which failed to reach consensus in our study) and turnaround time per case (which failed to reach high directional consensus).

Finally, it is worth noting some of the areas where there was either a lack of consensus among the panelists, or consensus toward a less optimistic perspective on the topic in question. There was a lack of consensus regarding whether AI would reduce the cost per case or the number of cases requiring pathologist review (after automated AI-based filtering of negative/normal cases), and whether it would increase patient satisfaction. There was also a lack of consensus regarding many regulatory aspects related to AI integration. For example, the panelists were uncertain whether it would become possible in the near future for approved developers to circumvent the requirement for individual approval of each new application, or whether legal and administrative barriers would be overcome regarding the use of de-identified images for research and education. There was general agreement that regulatory issues would still pose a challenge to the use of AI, even when the pathologist or another physician was the final decision-maker regarding diagnosis, prognostication, or treatment. There were differences of opinion surrounding several ethical aspects of AI integration, including whether AI outputs used for clinical decision-making would always need to be manually reviewed by a pathologist, or whether other healthcare professionals would start using AI tools to diagnose cases without the aid of a pathologist. There was uncertainty around whether the “black box” nature of many AI tools would cause pathologists to make diagnoses without enough clinical explainability, and a lack of consensus regarding whether pathologists would occasionally make diagnoses contrary to their own judgment because of AI software recommendations. There was also no consensus on whether AI would lead to de-skilling of pathologists (a subject which remains controversial within the pathology community^34^), or whether it would be possible to ensure that pathologists took full responsibility for double-checking and confirming AI-rendered diagnoses. Due to the current AI “translation gap” in pathology, there have been either no, or only a limited number of, studies evaluating the impact of AI tools on pathologist behavior,^25,26,59,68–71^ laboratory cost expenditures, medicolegal liability, or patient satisfaction. Therefore, the current lack of consensus among the panelists regarding the preceding topics is understandable and expected to be resolved as (1) more AI tools reach maturity and come to be evaluated in prospective clinical settings, and (2) more consideration is directed toward ensuring that AI models are integrated into pathology workflows in a manner that maximizes accuracy, time and cost efficiency, safety, transparency, accountability, and positive impact on patient outcomes. ^8,9,15,49,72^

Our study was subject to several limitations. Given the voluntary nature of participation and the substantial time commitment required to complete all three Delphi rounds, not all experts who were initially invited agreed to participate in the study, introducing the possibility of non-response bias. In addition, the proportion of female participants (16.7%) was much lower than the proportion of male participants. Although out of the 39 candidates originally invited to participate in this study 15 (38·9%) were women, possibly reflecting a relative underrepresentation of women in the pathology informatics and more specifically, the computational pathology/AI space, finally only four completed our survey. Only one of the participants was a cytopathologist (who also subspecialized in other areas of anatomic pathology), which may reflect the current underrepresentation of this subspecialty in computational pathology/AI due to the challenges of generating and storing z-stacked WSIs (which may resolve with the introduction of dedicated cytology whole-slide scanners in the near future).^42^ Also, as most of the participants were practicing in North America and Europe, the results of this study may reflect a predominantly North American/European perspective that differs from the perspectives of those practicing in other parts of the world. Lastly, as our study was targeted toward a specific respondent demographic, individuals from other stakeholder groups (such as pathology trainees, pathologists with little or no experience in computational pathology/AI, pathology laboratory technicians, computer scientists and others working in the computational pathology/AI space with non-medical backgrounds, physicians in other specialties, and patients) were not represented in our results. We anticipate all above issues to be sufficiently addressed in forthcoming studies, and that our survey, with its freely-available data collection forms, may serve as a model study for independent validation and extension.

We hope the results of this first systematic consensus study have provided a detailed vision of what pathology might look like in the year 2030, from the point of view of those with first-hand experience developing and evaluating clinical AI tools. Furthermore, we feel this study lays the groundwork for future follow-up studies in 3-5 years’ and 10 years’ time, assessing the evolution of expectations and challenges as the field of computational pathology progresses. Clearly, AI is expected to have a deep impact on the specialty of pathology, with several pathology AI applications anticipated to be in routine use by 2030, including some that will fully replace pathologists on specific tasks. The results of our study provide detailed insight into the current challenges, expectations, and perspectives surrounding the near to medium term role of AI in pathology, including timely and relevant information regarding how pathology care might be delivered in the future, assuming all regulatory and ethical questions are addressed.^48,73^ We expect these findings will be of broad interest to a wide variety of stakeholders, including pathologists, pathology trainees and laboratory technicians, hospital administrators, researchers in academia, industry, and government, patients, professional societies, and regulatory bodies.

## Data Availability

All data produced in the present study are available upon reasonable request to the authors

